# The risk of vertebral fractures in patients with differentiated thyroid cancer undergoing TSH suppression therapy

**DOI:** 10.64898/2025.12.01.25341205

**Authors:** Dilek Gogas Yavuz, Ceyda Dincer Yazan, Zeliha Hekimsoy, Kadriye Aydin, Naile Gokkaya, Canan Ersoy, Berna Imge Aydogan, Aysen Akalın, Omercan Topaloglu, Esra Nur Ademoglu Dilekci, Guven Baris Cansu, Bulent Can, Kaan Gungor, Mehtap Evran, Lezzan Keskin, Ziynet Alphan, Ozlem Turhan Iyidir, Levent Ozsari, Tayfun Galip, Gulsah Elbuken, Meral Mert, Zafer Pekkolay, Zeynep Canturk, Can Ilgın, Onur Bugdayci, Nilufer Ozdemir Kutbay, Goknur Yorulmaz, Ahmet Toygar Kalkan, Yasemin Aydogan Unsal, Adnan Yay, Baris Karagun, Evin Bozkur

## Abstract

**Backgound:** Suppression of thyroid-stimulating hormone (TSH) can adversely affect bone tissue in individuals with differentiated thyroid cancer (DTC).In a multi-center setting, this study aim to examine vertebral fractures (VF) and bone mineral density (BMD) in DTC patients receiving levothyroxine (LT4) suppression therapy.

**Methods:** Clinical measurements done in the last visit, laboratory and radiologic data extracted from patients files. Serum TSH concentrations were divided into four groups based on the American Thyroid Association’s guidelines for TSH suppression: 0.01 mu/L, 0.01-0.1, 0.1–0.5 mu/L, and >0.5 mu/L. Within the preceding year, the BMD measurements obtained in the lumbar and femoral regions with a DXA scanner were recorded. Lateral spinal X-rays obtained within the previous year were evaluated.

**Results:** The study comprised 990 people with DTC. The DXA showed 545 patients with normal BMD, 82 (8.2%) with osteoporosis, and 363 (36.6%). patients had osteopenia. The osteoporotic group exhibited older age (p = 0.001), lower BMI (p = 0.001), and comparable TSH levels compare to the osteopenia and normal BMD groups. In 476 lateral vertebral x-rays, 200 (42%), 131 (27.5%) were significant or severe VF. The severe TSH suppression group had a greater fracture rate than the moderate, mild, and non-suppressed groups(p=0.013). VF patients were older (p = 0.001) and had lower femur neck and L1-L4 BMD. Disease duration, FT4 level, and femur neck BMD affected fracture risk (p = 0.001, pseudo R2: 0.028). Age, femur neck BMD, L1-L4 BMD, and osteocalcin levels were linked with osteoporosis in a multivariate analysis (p = 0.001, pseudo R2: 0.427). TSH subgroups had similar femur neck (p = 0.61) and L1-L4 BMD (p = 0.89).

**Conclusion:** Findings suggest that DTC patients undergoing TSH suppression have a significant risk of VF. Assessment of VF should be integrated into the routine care of DTC survivors to ensure long-term skeletal safety.

## 1. Introduction

Thyroid Stimulating Hormone (TSH) suppression treatment is usually recommended for differentiated thyroid cancer (DTC) patients following thyroidectomy in order to reduce the risk of recurrence and mortality for those at intermediate and high risk, as determined by their postoperative response status. [1,2].

Recent evidence indicates that TSH over-suppression is a common clinical concern potentially associated with bone complications[3].

The effects of iatrogenic hyperthyroidism on BMD and fracture risk are controversial in DTC patients.The studies on bone mineral density (BMD) and fracture risk involved a limited cohort of DTC patients, yielding inconsistent results due to gender differences and variability within the study populations. According to a case-control study that included both a control group and patients with thyroid cancer, U.S. Veterans with thyroid cancer had a higher risk of osteoporosis than controls, but not fractures.Lower TSH levels, female sex, advanced age, and androgen use are associated with osteoporosis. TSH levels showed no correlation with fracture incidence. [4].

In a meta-analysis from observational studies suggests that postmenopausal women treated with TSH suppression therapy after total thyroidectomy are at risk for lower BMD than premenopausal women [5].An other meta-analysis reported that stringent TSH suppression (TSH <0.10 mIU/L) after thyroidectomy had deleterious effects on the BMD of the lumbar spine in postmenopausal DCT patients compared with controls [6].

In a Korean study with thyroid cancer patients, osteoporosis was higher in thyroid cancer patient group but fracture status was comparable with controls [7].A small cohort of DTC patients was shown to have no increased fracture risk [8].Long-term suppression with levothyroxine in men with DTC does not appear to have a negative effect on bone mineral density or increase the incidence of vertebral fractures [9].

A European study indicated a high prevalence of VFs among women with DTC undergoing long-term, suppressive L-T4 treatment [10]. A recent study indicated that the prevalence of VF was significantly higher in the Papillary thyroid carcinoma cohort (44.1%) compared to healthy controls in the Japan differentiated thyroid carcinoma group [11].

The main purpose of this study was to determine the frequency of vertebral fractures, clinical determinants of vertebral fractures, and bone mineral density measurements in DTC patients receiving levothyroxine (LT4) suppression treatment after total thyroidectomy with or without radioiodine ablation therapy in a multicenter, nationwide setting.

## 2. Materials and Methods

This cross-sectional study was conducted at 21 medical centers in 12 cities (Istanbul, Kocaeli, Manisa, Bursa, Eskisehir, Malatya, Diyarbakir, Adana, Usak, Batman, Bolu, Tekirdag) in different regions of Turkey. The study was approved by the Local Ethics Committee (09.2017.359) and conducted in compliance with the Declaration of Helsinki. Laboratory values from the most recent visit and BMD measurements, vertebral X rays taken within the previous year were obtained from files, and data were collected between the years 2018 and 2020.

### 2.1. Patients selection

This research involved 990 patients diagnosed with differentiated thyroid cancer (DTC) who underwent either total or subtotal thyroidectomy, with or without subsequent radioiodine ablation, and were observed for a minimum duration of one year while undergoing levothyroxine suppression therapy. The study excluded participants with central hypothyroidism, inflammatory bowel disease, malabsorption syndrome, gastric bypass surgery, other malignancies, bone metastases, and medullary or anaplastic thyroid cancer. Individuals receiving medications that influence bone metabolism, such as glucocorticoids antiepileptic drugs and hormone replacement were excluded. Individuals currently receiving tri-iodothyronine (T3) therapy were excluded from the study.

### 2.2. Clinical evaluation

Clinical data including age, sex, disease duration, surgical type (total or subtotal), radioactive iodine treatment, and comorbid diseases such as hypertension, diabetes mellitus, cerebrovascular disease, coronary artery disease, atrial fibrillation, malabsorption, inflammatory bowel disease, and gluten enteropathy were extracted from medical records.

Data from pathology reports was extracted from files. The daily doses of LT4 were documented. The quantity of LT4 per kilogram was determined.

Body weight and height were measured, and the BMI was calculated as kg/m2 using the BMI formula. After 15 minutes of rest, the systolic and diastolic blood pressures and heart rate were measured.

### 2.3. Laboratory evaluation

Serum TSH, FT4, serum Thyroglobulin (Tg), and anti-Thyroglobulin (Anti-Tg)levels have been recorded from patients’ files within three months. Mean level of the current, 6 months ago and 1 year ago TSH levels were calculated for each patient. The third-generation chemiluminescence immunoassay method was employed to quantify thyroid hormones in hospital laboratories, where patient monitoring was also conducted. The laboratory reference ranges for TSH and FT4 were similar. The reference range for TSH is 0.34-5.60 mU/L, while FT4 ranges from 0.61 to 1.12 ng/dl (0.144-0.26 pmol/L). The minimum detectable level of TSH was 0.01 mIU/L. Venous blood samples for thyroid function analysis were collected after an 8-hour fast, prior to the ingestion of the morning pill.

Levels of serum calcium, phosphorus, parathormone, 25-hydroxyvitamin D3, osteocalcin, and c-telopeptide were documented in patient files during the latest visit. 25(OH)D levels were assessed utilizing the Beckman Coulter paramagnetic particle chemiluminescence immunoassay technique. The Cobas 6000 immunoassay (Roche) was employed to quantify intact parathormone levels, with a reference range of 15-65 ng/L. Inorganic phosphorus (reference range: 2.6-4.5 mg/dl) and calcium (reference range: 8.8-10.6 mg/dl) were measured photometrically (Beckman Coulter, USA), whereas osteocalcin (3-13 mcg/L) and c-telopeptide (0.02-0.57 ng/ml) were measured by chemiluminescence immunoassay.

The ATA management guidelines for adult thyroid cancer patients recommend maintaining target TSH levels at 0.1 mU/L for high-risk individuals and between 0.1-0.5 mU/L for those at intermediate risk (1).Patients’ TSH levels were classified as severely suppressed (TSH 0.01 mU/L), moderately suppressed (TSH: 0.01-0.1 mU/L), mildly suppressed (TSH: 0.1-0.5 mU/ L), or non-suppressed (TSH>0.5 mU/L) according to ranges defined in ATA guideline. Patients were also classified based on their achievement of target thyroid-stimulating hormone (TSH) levels, which were adjusted for characteristics such as disease duration, risk and remission. This classification differentiated between patients who achieved the target levels and those who did not.

### 2.4. Bone mineral density

BMD was assessed by dual-energy X-ray absorptiometry (DXA) and measured in the lumbar spine (L1-L4) in the anteroposterior (AP) projection, as well as the three sites of the right hip (femoral neck, using a Lunar DPX-L) within one year in all centers. All of the BMD reports were evaluated for accuracy of shooting technique. Sixty-eight percent of repeated measurements were statistically within one standard deviation (0.012 g/cm3).

Osteopenia and osteoporosis were classified in accordance with the standards established by the World Health Organization, which rely on T and Z scores. Osteopenia is defined as a T score lying within the range of -1 standard deviation (SD) to -2.5 SD for either the lumbar anterior-posterior (AP) or femoral neck. Osteoporosis is characterized by a bone mineral density (BMD) T score, evaluated using dual-energy X-ray absorptiometry (DXA), that falls at or below -2.5 standard deviations (SD) from the mean at either the lumbar spine or femoral neck. This diagnostic criterion applies exclusively to postmenopausal women and males who are 50 years of age or older. It was defined as a BMD Z score on DXA at the lumbar spine or femoral neck less than or equal to -2 SD for premenopausal women and males under the age of 50 as low bone mineral density according to chronological age and included within osteoporotic group.

### 2.5. Vertebral fracture evaluation

476 of the patients had vertebral x-ray. Vertebral fractures were assessed by lateral thoracolumbar X-ray radiograms, which were performed independently by two specialists. The semi-quantitative technique developed by Genant et al. [12].was applied. The grading system divided fractures into four categories: normal (grade 0), mild (grade 1), which involved a reduction of approximately 20-25% in anterior, middle, and/or posterior height and a reduction of area by 10-20%, moderate (grade 2), which involved a reduction of approximately 25-40% in any height and a reduction of area by 20-40%, and severe (grade 3), which involved a reduction of approximately 40% in any height and area. Vertebral radiograms were independently evaluated by two blinded specialists (O.B/ and D.G.Y).

### 2.6. Statistical Analysis

The distribution of the data was evaluated by Shapiro-Wilk Test. One Way Anova test was employed for comparing the data demonstrating normal distribution among the three groups. The Kruskal–Wallis ANOVA test was used to compare variables that did not have a normal distribution among the three groups. The Fisher Exact Test examined the differences between categorical data sets. Post-hoc comparisons of the variables found meaningful after the ANOVA and Kruskal–the Dunn test analysed Wallis tests. Descriptive statistics of data for numerical variables: average, standard deviation,and categorical variables were given as frequency.Bonferroni correction was applied in multiple group analysis. The significance level for subgroup analysis was p<0.0167 for three parameters and p<0.0083 for four parameters.All analyses were executed using Stata 15.1 software (StataCorp, Texas 77845 USA).

## 3. Results

### 3.1. Clinical characteristics of the patients

There were 990 DTC patients enrolled (F/M:829/161), with a mean age of 51.4±11.5 years and a mean disease duration of 5.2±4.4 years. Average daily intake of LT4 was 131±37.5 mcg/day. Table 1 presents the clinical and laboratory features of the study group categorized by gender. The mean TSH level within one year of the overall study group was 1.94±8.02 mU/L. The mean femur neck BMD was 0.930.18 g/cm 2, while the L1-L4 BMD was 1.070.204 g/cm 2. Women were younger (p=0.037) and had a higher BMI (p<0.001) than men. Women had a lower daily total LT4 dosage (p<0.001), LT4 dose/kg (p<0.001), BMD at L1-L4 (p<0.001), and femur neck BMD (p<0.001), than men. TSH and fT4 levels were similar according to gender. Men had greater serum calcium (p<0.001) and 25(OH)D levels (p=0.001) than women.

**Table 1.** Clinical and laboratory characteristics of patients with differentiated thyroid cancer.

|  | **Total (n=990)** | **Men (n=161)** | **Women (n=829)** | **P\*** |
| --- | --- | --- | --- | --- |
| **Age (years)** | 51.4±11.5 | 53.3±12.5 | 51.07±11.2 | 0.037 |
| **Duration of disease (years)** | 5.2±4.4 | 4.85±4.64 | 5.2±4.4 | 0.13 |
| **BMI (kg/m<sup>2</sup>)** | 30.5±5.69 | 28.9±4.22 | 30.8±5.89 | <0.001 |
| **SBP (mmHg)** | 125.6±19 | 129.2±19.2 | 125±18.9 | 0.018 |
| **DBP (mm/Hg)** | 77.7±11.89 | 76.2±19.2 | 77.4±11.7 | 0.12 |
| **Pulse (per minute)** | 79.6±10.2 | 82±12.3 | 79.2±9.83 | 0.051 |
| **LT4 dose mcg/kg** | 1.69±0.47 | 1.67±0.48 | 1.79±0.45 | <0.001 |
| **LT4 daily dosage (mcg/day)** | 131±37.5 | 152.2±41.3 | 126.9±35.3 | <0.001 |
| **Femur neck BMD (g/cm<sup>2</sup>)** | 0.930±0.18 | 1.000±0.18 | 0.925±0.18 | <0.001 |
| **Femur neck T score** | -0.31±1.24 | -0.23±1.32 | -0.3±1.23 | 0.41 |
| **Femur neck z score** | 0.29±1.14 | 0.38±1.35 | 0.28±1.105 | 0.17 |
| **L1-L4 BMD(g/cm<sup>2</sup>)** | 1.070±0.204 | 1.130±0.22 | 1.060±0.19 | <0.001 |
| **L1-L4 T score** | -0.44±1.63 | -0.07±1.73 | -0.51±1.60 | 0.0038 |
| **L1-L4 Z score** | -0.005±1.43 | 0.18±1.75 | -0.04±1.36 | 0.23 |
| **TSH (mU/L)** | 1.94±8.02 | 1.73±4.43 | 1.99±8.54 | 0.71 |
| **FT4 (ng/dl)** | 3.38±4.91 | 3.3±4.87 | 3.4±4.92 | 0.77 |
| **Calcium (mg/dl)** | 9.47±0.59 | 9.6±0.52 | 9.45±0.60 | <0.001 |
| **Phosphorus (mg/dl)** | 3.59±0.72 | 3.3±0.65 | 3.65±0.72 | <0.001 |
| **Parathormone (ng/dl)** | 52.5±37.5 | 53.1±36.8 | 51.8±33.5 | 0.66 |
| **25-OH-D (ng/dl)** | 22.45±12.9 | 24.8±11.7 | 22.0±13.09 | 0.001 |
| **Osteocalcin(ng/ml )** | 5.11±4.29 | 4.8±3.5 | 5.1±4.4 | 0.81 |
| **C-telopeptide (ng/ml)** | 0.86±5.85 | 0.75±0.6 | 0.88±0.75 | 0.90 |

### 3.2. Assessment of Bone Mineral Density

BMD evaluation revealed 545 patients (55.2%) have normal BMD, 82 individuals (8.2%) have osteoporosis while 363 patients (36.6%) had osteopenia.

The patients with normal BMD were younger than the osteopenic (p<0.001) and osteoporotic patients (p=0.005). The BMI of the osteoporotic/low bone mass group was significantly lower than that of the osteopenic (p<0.001) and the normal group (p<0.001). The osteoporotic group had a higher serum PTH than the osteopenic and normal BMD groups (p<0.001). Serum 25-OH D levels of the groups were similar. Patients with osteoporosis had higher osteocalcin levels than the normal group (p<0.001). Laboratory and clinical characteristics of the osteoporotic,osteopenic and normal BMD group’s are shown in Table 2.

**Table 2.**
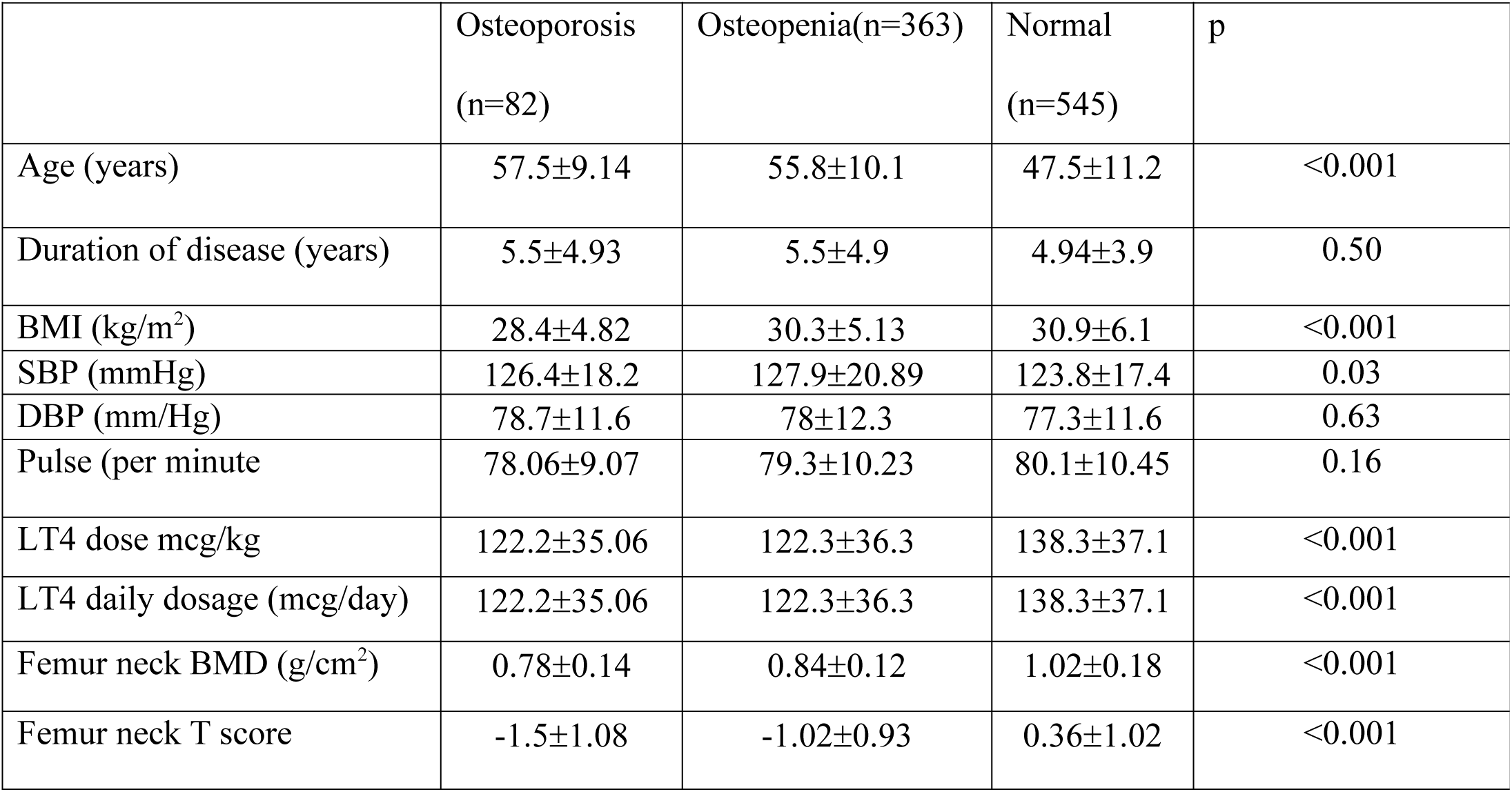
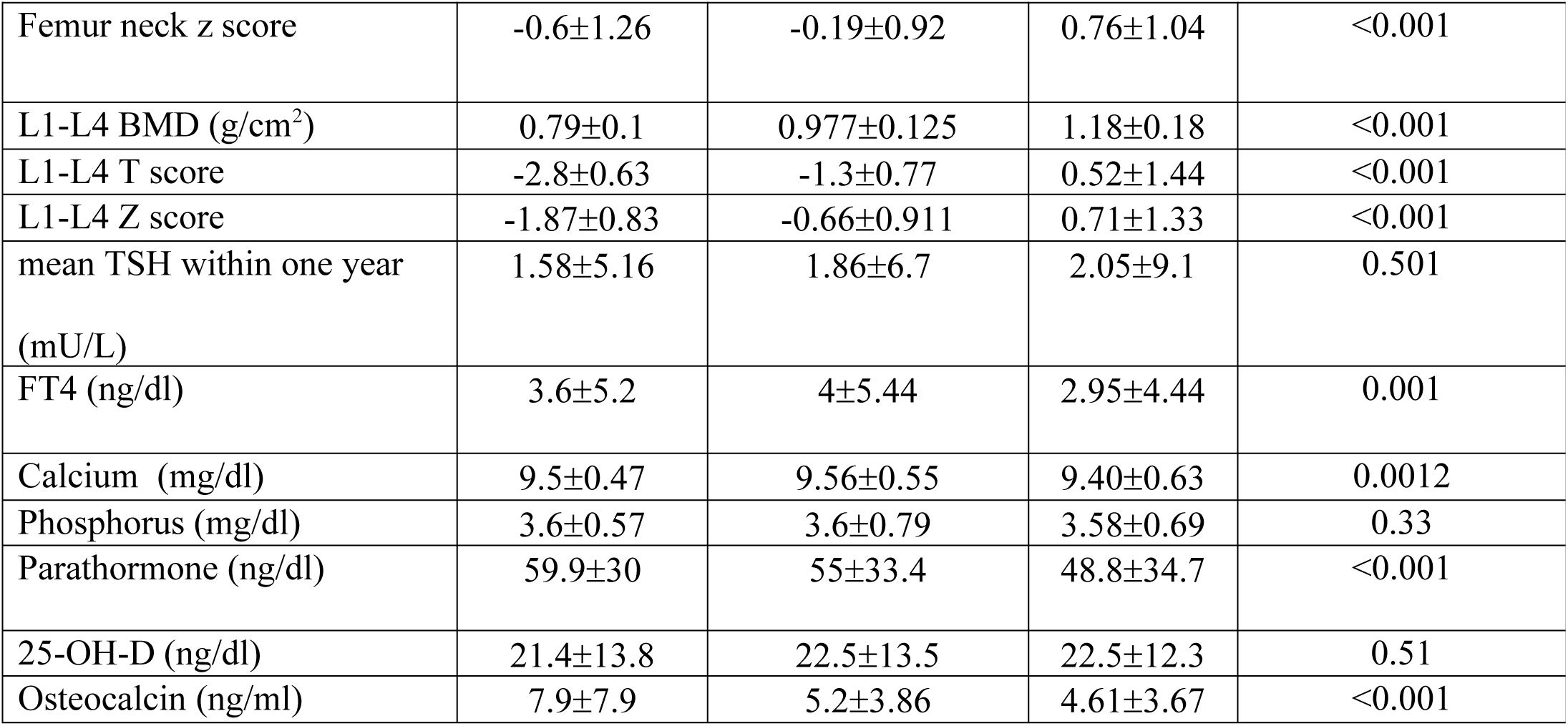
Clinical and laboratory values of patients according to osteoporosis status.

|  | Osteoporosis<br>(n=82) | Osteopenia(n=363) | Normal<br>(n=545) | p |
| --- | --- | --- | --- | --- |
| Age (years) | 57.5±9.14 | 55.8±10.1 | 47.5±11.2 | <0.001 |
| Duration of disease (years) | 5.5±4.93 | 5.5±4.9 | 4.94±3.9 | 0.50 |
| BMI (kg/m<sup>2</sup>) | 28.4±4.82 | 30.3±5.13 | 30.9±6.1 | <0.001 |
| SBP (mmHg) | 126.4±18.2 | 127.9±20.89 | 123.8±17.4 | 0.03 |
| DBP (mm/Hg) | 78.7±11.6 | 78±12.3 | 77.3±11.6 | 0.63 |
| Pulse (per minute) | 78.06±9.07 | 79.3±10.23 | 80.1±10.45 | 0.16 |
| LT4 dose mcg/kg | 122.2±35.06 | 122.3±36.3 | 138.3±37.1 | <0.001 |
| LT4 daily dosage (mcg/day) | 122.2±35.06 | 122.3±36.3 | 138.3±37.1 | <0.001 |
| Femur neck BMD (g/cm<sup>2</sup>) | 0.78±0.14 | 0.84±0.12 | 1.02±0.18 | <0.001 |
| Femur neck T score | -1.5±1.08 | -1.02±0.93 | 0.36±1.02 | <0.001 |
| Femur neck z score | -0.6±1.26 | -0.19±0.92 | 0.76±1.04 | <0.001 |
| L1-L4 BMD (g/cm<sup>2</sup>) | 0.79±0.1 | 0.977±0.125 | 1.18±0.18 | <0.001 |
| L1-L4 T score | -2.8±0.63 | -1.3±0.77 | 0.52±1.44 | <0.001 |
| L1-L4 Z score | -1.87±0.83 | -0.66±0.911 | 0.71±1.33 | <0.001 |
| mean TSH within one year<br>(mU/L) | 1.58±5.16 | 1.86±6.7 | 2.05±9.1 | 0.501 |
| FT4 (ng/dl) | 3.6±5.2 | 4±5.44 | 2.95±4.44 | 0.001 |
| Calcium (mg/dl) | 9.5±0.47 | 9.56±0.55 | 9.40±0.63 | 0.0012 |
| Phosphorus (mg/dl) | 3.6±0.57 | 3.6±0.79 | 3.58±0.69 | 0.33 |
| Parathormone (ng/dl) | 59.9±30 | 55±33.4 | 48.8±34.7 | <0.001 |
| 25-OH-D (ng/dl) | 21.4±13.8 | 22.5±13.5 | 22.5±12.3 | 0.51 |
| Osteocalcin (ng/ml) | 7.9±7.9 | 5.2±3.86 | 4.61±3.67 | <0.001 |

### 3.3. Assessment vertebral fracture

A total of 476 patients were evaluated using the lateral spinal X-ray. A total of 200 patients (42.0%) displayed a minimum of a grade 1 fracture, and 131 (27.5%) had moderate / severe vertebral fractures during the assessment of spinal x-rays.

Seventeen of the 131 patients with moderate-severe fracture were premenopausal or male under 50 years. The average number of vertebral fractures was 5.43. A total of 193 patients exhibited the presence of at least one thoracic vertebral fracture, whereas 87 patients displayed the occurrence of at least one lumbar vertebral fracture. Table 3 displays the clinical and laboratory data of patients according to vertebral fracture status. The age of the individuals with fractures was significantly higher (p<0.001) compared to the non-fractured group. There were no significant differences seen in the daily dosage (p=0.1) and dose per kg (p=0.13) between the groups. The femur neck bone mineral density (BMD) of the fracture group was significantly lower compared to patients without fractures (p=0.0001). Additionally, the L1-L4 BMD of the fracture group was also significantly lower than that of patients without fractures (p=0.0001). Patients with fractures had higher TSH levels (p=0.0088) than those without fractures.

**Table 3:**
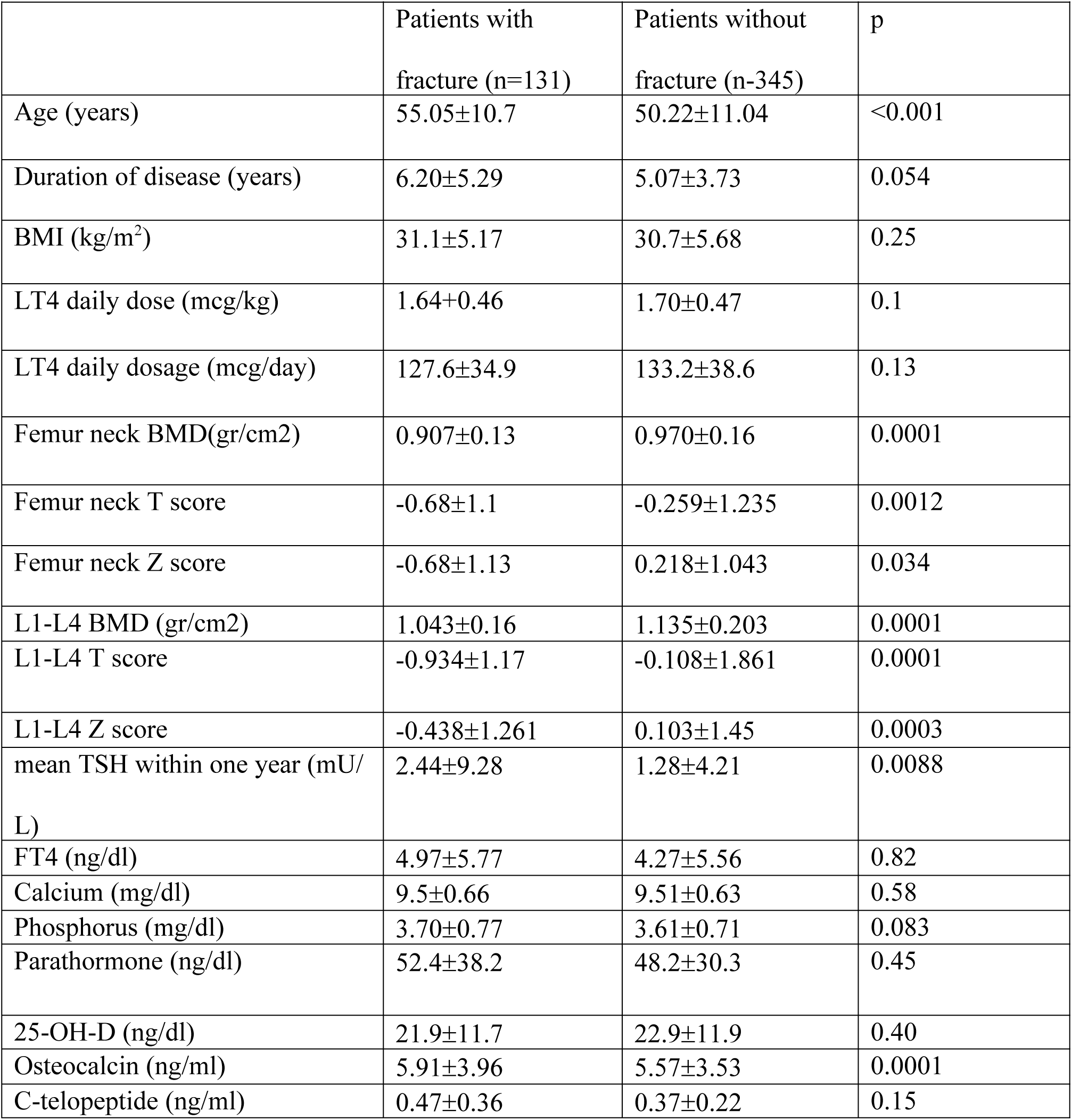
DTC patients’ clinical and laboratory characteristics by status of vertebral fractures.

According to the TSH suppression,patients in the severely, moderately, and midlysupressed and nonsupressed groups experienced at least one vertebral fracture at rates of 44% (n=8), 15.8% (n=49), and 15.6% (n=43 ), 25.8% (n= 99 ) respectively. The fracture rate in the severe suppression group was significantly greater than in the moderate, mild, and nonsuppressed groups (p = 0.013).

Five patients (27.7%) in the severe suppression group, 29 (9.35%) in the moderate suppression group, 29 (10.5%) in the mild suppression group, and 68 (16.2%) in the non-suppressed group all experienced grade 2 or 3 vertebral fractures.

### 3.4. Assessment of patients according to TSH levels

Table 4 presents the clinical and laboratory characteristics of patients according to TSH suppression level. Thirty-three percent of patients had TSH levels below 0.1 mIU/L. The duration of the illness varied between groups. The duration of the disease was longer in groups with mild suppression (p=0.006, p<0.001) than in groups with moderate or severe suppression (p<0.001).

**Table 4.** DTC patients’ clinical and laboratory characteristics according to TSH suppression.

|  | TSH<0.01<br>(n=18) | TSH 0.01-0.1<br>(n=310) | TSH 0.1-0.5<br>(n=275) | TSH>0.5<br>(n=387) | p |
| --- | --- | --- | --- | --- | --- |
| Age (years) | 48.5±12 | 50.3±10.7 | 51.8±11.8 | 52.1±11.86 | 0.11 |
| Duration of disease years) | 3.61±3.95 | 4.16±3.65 | 5.58±4.22 | 5.84±5.02 | <0.001 |
| BMI (kg/m<sup>2</sup>) | 29.4±3.88 | 29.5±5.31 | 31.1±5.87 | 30.9±5.85 | 0.014 |
| SBP (mmHg) | 124.5±15.9 | 124±19.06 | 123±18.1 | 128.3±19.4 | 0.018 |
| DBP (mm/Hg) | 79.8±9.12 | 77.6±12.1 | 75.2±11.1 | 79.4±12 | 0.003 |
| Pulse (per minute) | 81.6±4.94 | 80.4±9.67 | 79.4±10.5 | 78.9±10.4 | 0.14 |
| LT4 dose mcg/kg | 1.88±0.81 | 1.82±0.48 | 1.65±0.43 | 1.60±0.45 | <0.001 |
| LT4 daily dosage (mcg/day) | 134±56.9 | 138±36.8 | 130.1±34.8 | 125.9±38 | <0.001 |
| Femur neck BMD (g/cm<sup>2</sup>) | 0.880±0.14 | 0.940±0.16 | 0.951±0.18 | 0.929±0.205 | 0.61 |
| Femur neck T score | -0.81±1.03 | -0.307±1.21 | -0.16±1.35 | -0.39±1.2 | 0.21 |
| Femur neck z score | -0.13±0.75 | 0.289±1.09 | 0.44±1.21 | 0.22±1.15 | 0.16 |
| L1-L4 BMD (g/cm<sup>2</sup>) | 1.080±0.14 | 1.075±0.21 | 1.080±0.206 | 1.07±0.19 | 0.89 |
| L1-L4 T score | -0.62±1.08 | -0.43±1.58 | -0.44±1.51 | -0.44±1.78 | 0.93 |
| L1-L4 Z score | -0.22±1.1 | -0.035±1.5 | 0.03±1.40 | -0.0003±1.4 | 0.83 |
| TSH (mU/L) | 0.005±0.002 | 0.035±0.025 | 0.24±0.109 | 4.78±12.3 | <0.001 |
| FT4 (ng/dl) | 10.1±8.15 | 3.18±4.86 | 3.18±4.78 | 3.45±4.72 | <0.001 |
| Calcium (mg/dl) | 9.42±0.65 | 9.44±0.58 | 9.49±0.58 | 9.49±0.614 | 0.48 |
| Phosphorus (mg/dl) | 4.16±0.66 | 3.53±0.72 | 3.58±0.72 | ±0.71 | 0.002 |
| Parathormone (ng/dl) | 39.2±17.5 | 52.4±34.5 | 52.9±38.7 | 51.7±30.4 | 0.42 |
| 25-OH-D (ng/dl) | 23.7±11.2 | 22.07±14.2 | 22.6±13.6 | 22.6±11.3 | 0.34 |
| Osteocalcin (ng/ml) | 6.56±4.51 | 5.77±4.61 | 4.49±3.28 | 4.94±4.58 | 0.064 |
| C-telopeptide (ng/ml) | 0.42±0.31 | 0.43±0.35 | 0.31±0.12 | 0.305±0.28 | 0.0064 |
| Data were presented as mean standard deviation |  |  |  |  |  |

The duration of the disease was longer in the non-suppressed group than in the severe suppression group (p<0.001). The moderate suppression group’s BMI was lower than that of the non-suppressed group (p=0.0056) and mild suppression group (p=0.0061). Daily doses of levothyroxine varied by group (p<0.001). The moderately suppressed group received a greater daily dose of levothyroxine than the non-suppressed groups (p<0.001). The severe suppression group had greater serum phosphorus levels than the moderate suppression (p<0.001), mild suppression (p<0.001), and non-suppressed (p=0.0025) groups. The moderately suppressed group had a higher amount of serum c-telopeptide than the non-suppressed group (p<0.001). Based on suppression levels, the patient’s femur neck BMD and L1-L4 BMD levels were comparable among groups.

### 3.5. The assessment of patients according to the menopausal state

Fivehundredandfivewomenwerecategorized as postmenopausal. Table 5 showsthelaboratoryanalysis of participantsgroupedbytheirmenopausestatus.Premenopausal and postmenopausal patients had similar TSH levels. L1-L4 and femur neck BMD measurements in the postmenopausal group were lower than those in the premenopausal group (p<0.001). Premenopausal women and men had lower calcium and 25(OH)D levels than postmenopausal women and men (p<0.001).

**Table 5.**
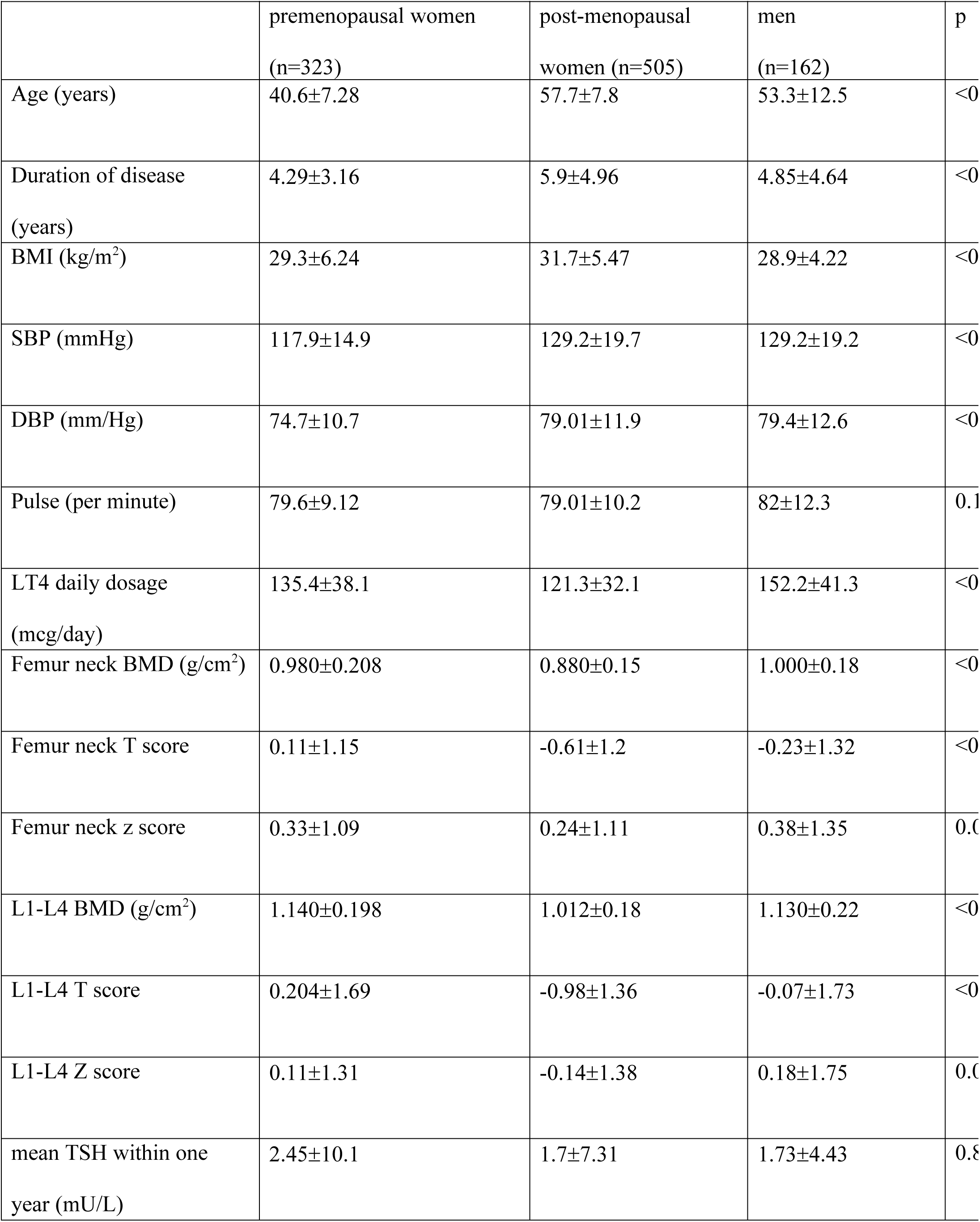
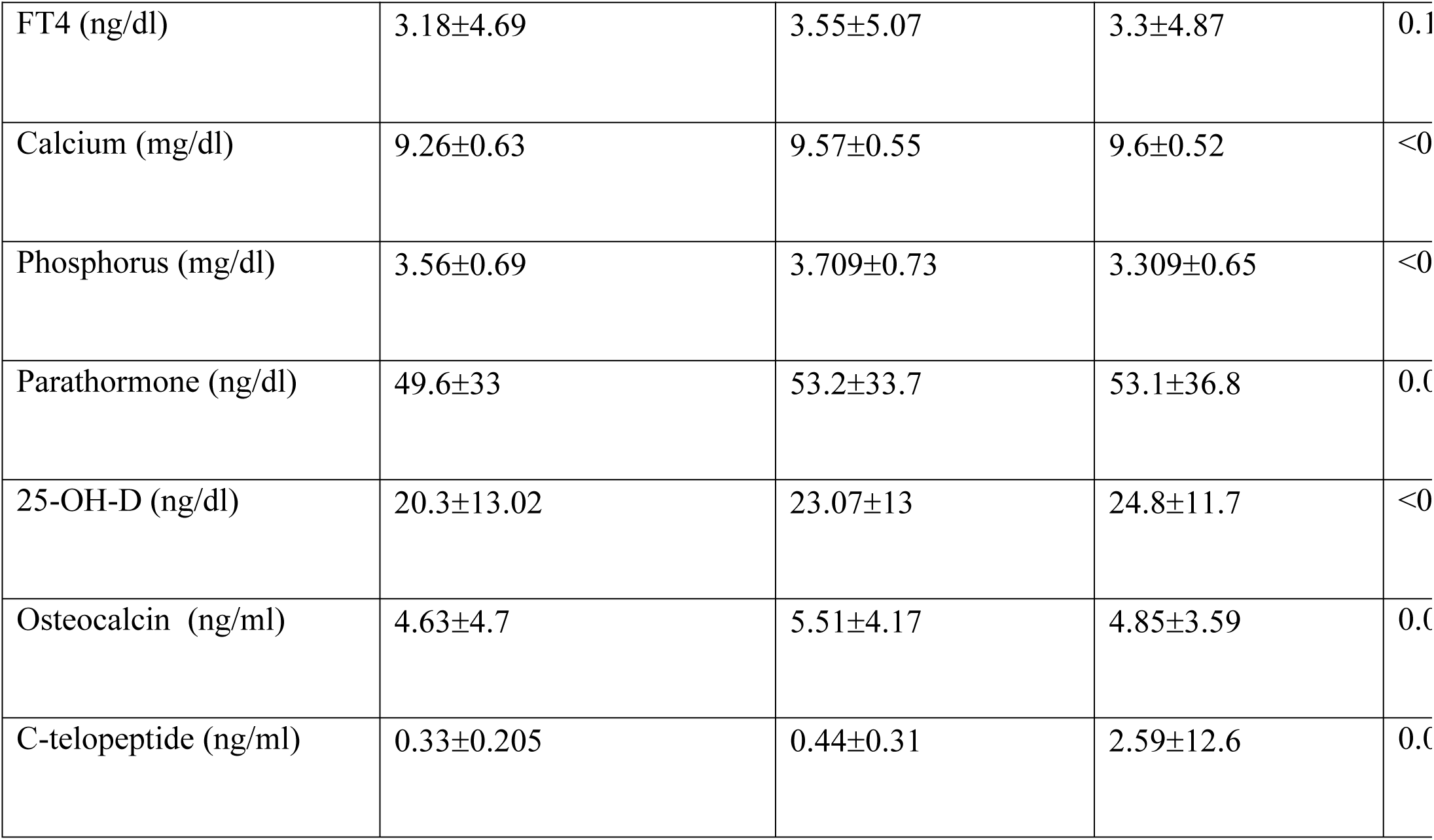
Clinical and laboratory characteristics according to sex and menopausal status.

*Tablo 5 Clinical and laboratory characteristics according to sex and menopausal status*
|  | premenopausal women<br>(n=323) | post-menopausal<br>women (n=505) | men<br>(n=162) | p |
| --- | --- | --- | --- | --- |
| Age (years) | 40.6±7.28 | 57.7±7.8 | 53.3±12.5 | <0 |
| Duration of disease<br>(years) | 4.29±3.16 | 5.9±4.96 | 4.85±4.64 | <0 |
| BMI (kg/m<sup>2</sup>) | 29.3±6.24 | 31.7±5.47 | 28.9±4.22 | <0 |
| SBP (mmHg) | 117.9±14.9 | 129.2±19.7 | 129.2±19.2 | <0 |
| DBP (mm/Hg) | 74.7±10.7 | 79.01±11.9 | 79.4±12.6 | <0 |
| Pulse (per minute) | 79.6±9.12 | 79.01±10.2 | 82±12.3 | 0.1 |
| LT4 daily dosage<br>(mcg/day) | 135.4±38.1 | 121.3±32.1 | 152.2±41.3 | <0 |
| Femur neck BMD (g/cm<sup>2</sup>) | 0.980±0.208 | 0.880±0.15 | 1.000±0.18 | <0 |
| Femur neck T score | 0.11±1.15 | -0.61±1.2 | -0.23±1.32 | <0 |
| Femur neck z score | 0.33±1.09 | 0.24±1.11 | 0.38±1.35 | 0.0 |
| L1-L4 BMD (g/cm<sup>2</sup>) | 1.140±0.198 | 1.012±0.18 | 1.130±0.22 | <0 |
| L1-L4 T score | 0.204±1.69 | -0.98±1.36 | -0.07±1.73 | <0 |
| L1-L4 Z score | 0.11±1.31 | -0.14±1.38 | 0.18±1.75 | 0.0 |
| mean TSH within one<br>year (mU/L) | 2.45±10.1 | 1.7±7.31 | 1.73±4.43 | 0.8 |
| FT4 (ng/dl) | 3.18±4.69 | 3.55±5.07 | 3.3±4.87 | 0.1 |
| Calcium (mg/dl) | 9.26±0.63 | 9.57±0.55 | 9.6±0.52 | <0 |
| Phosphorus (mg/dl) | 3.56±0.69 | 3.709±0.73 | 3.309±0.65 | <0 |
| Parathormone (ng/dl) | 49.6±33 | 53.2±33.7 | 53.1±36.8 | 0.0 |
| 25-OH-D (ng/dl) | 20.3±13.02 | 23.07±13 | 24.8±11.7 | <0 |
| Osteocalcin (ng/ml) | 4.63±4.7 | 5.51±4.17 | 4.85±3.59 | 0.0 |
| C-telopeptide (ng/ml) | 0.33±0.205 | 0.44±0.31 | 2.59±12.6 | 0.0 |

The body mass index (BMI) of the postmenopausal group exhibited a statistically significant increase compared to both the premenopausal women and males (p<0.001). The systolic blood pressure (SBP) (p < 0.001) and diastolic blood pressure (DBP) (p < 0.001) of the premenopausal group exhibited a statistically significant decrease when compared to both postmenopausal women and males.

No significant association was seen between TSH levels and femur and L1-L4 BMD levels in premenopausal (p=0.53, p=0.38), postmenopausal women (p=0.6, p=0.7), and men (p=0.59, p=0.5). There was no statistically significant link observed between the length of the disease and the levels of bone mineral density (BMD) in the femur and L1-L4 vertebrae (p = 0.11, p = 0.65) in all groups.

### 3.6. Analysis of osteoporotic and fracture status variables

In univariate analysis, age, sex, disease duration, mean daily LT4 dose/kg, femur neck BMD, L1-L4 BMD, c-telopeptide, and osteocalcin affected osteoporotic status. In multivariate analysis, age, femur neck BMD, L1-L4 BMD, and osteocalcin levels predicted osteoporosis (p<0.001, pseudoR^2^: 0.427).

The factors affecting the fracture status were age, sex, duration of disease, mean daily levothyroxine dose/kg, femur neck BMD, L1-L4 BMD, and FT4 levels in univariate analysis. In multivariate analysis, disease duration, FT4, and femur neck BMD were found to be predictive factors. (p<0.001, pseudo R^2^: 0.028).The multivariate analysis results were shown in table 6.

**Table 6.**
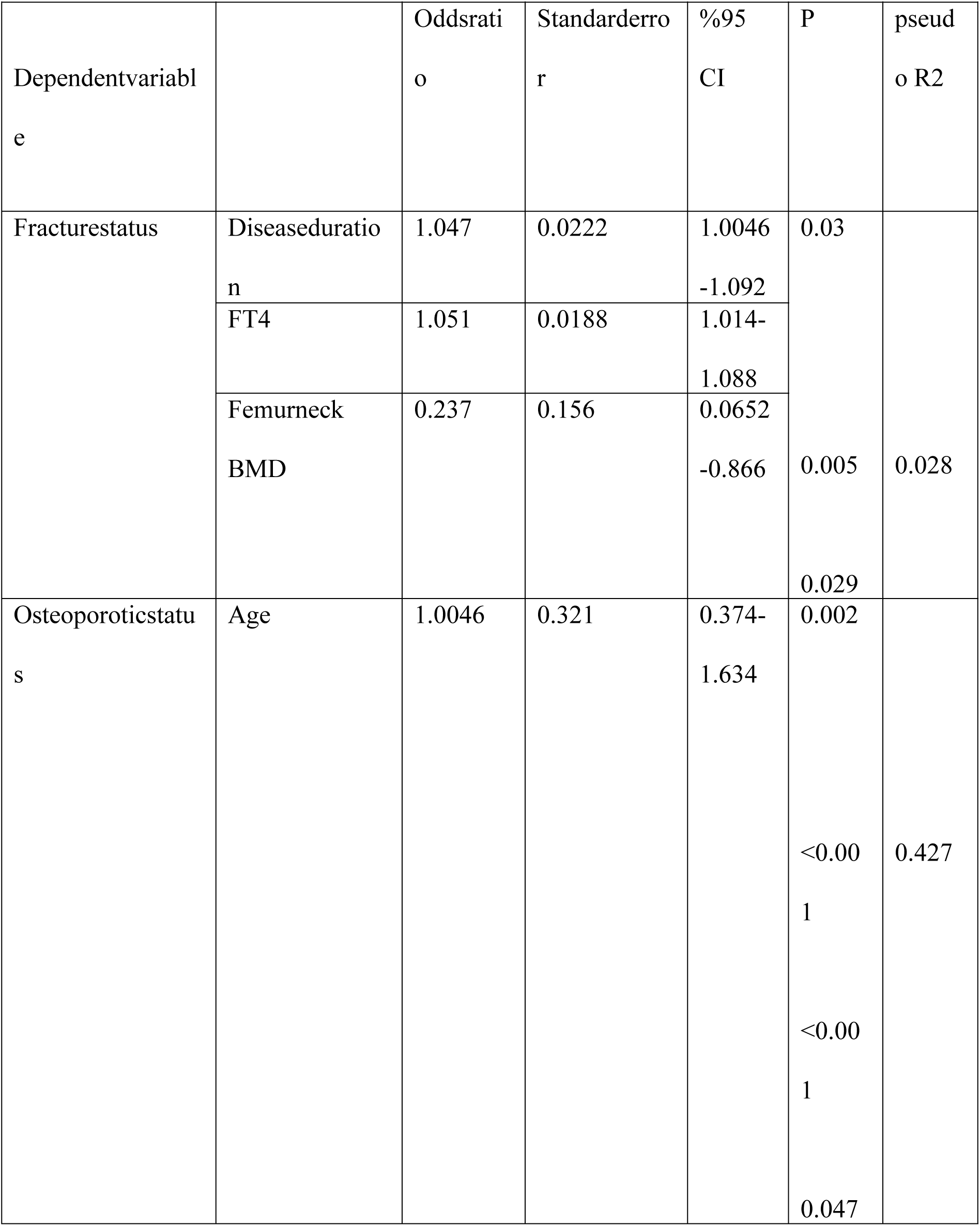
Multivariate analysis of osteoporotic and fracture statusin DTC patients.

## 4. Discussion

In the present study we reported42.0% of the thyroid cancer patients under LT4 treatment exhibited vertebral fracturesand 27.5%of them had moderate / severe vertebral fractures.Predicting factors were found to be disease duration, FT4 level, and femur neck BMD.

The VF rates we observed are in accordance with earlier European research that found that patients with DTC had greater VF rates than the general population.

Mazziotti et al. reported a 28.5% prevalence of radiological vertebral fractures, with a fracture prevalence of 44.6% in the TSH <0.5 mIU/L subgroup among 179 women undergoing TSH suppressive therapy for thyroid carcinoma. [10].A small study from Japan revealed that the prevalence of VFs was significantly higher in the PTC group (44.1%) compared to the control group (16.3%). [11].

Contrary studies have reported no increased risk of vertebral fractures under long-term LT4 suppression in patients with differentiated thyroid cancer, both men and women. [8,9].

We observed osteoporosis in 8.2% and osteopenia in 36.6% of the study group which is compatible with literature. In a study enrolled in patients with DTC, osteoporosis was observed in 7.3% of patients, which was more frequent according to control group (5.3%) [4]. The impact of TSH suppression treatment on bone mineral density in patients with differentiated thyroid cancer remains unclear, with conflicting results reported in the literature.Most cross-sectional studies in the literature report no significant change in BMD levels in premenopausal women compared to the control group [8, 14–20].However, two meta-analyses indicate detrimental effects on BMD levels in postmenopausal women and men [5–6].

A large-scale longitudinal investigation found that women over 50 year sold under going TSH suppression treatment had lower bone mineral density than women with thyroid cancer who had normal TSH levels [21]. Cross-sectional studies have also produced contradictory results [22–26].A study observed no difference in BMD levels between treatment and control groups after 7.3 years of TSH suppression medications in DTC patients [22].

After 12.2 years of TSH suppression therapy, Chinese women reported a decline in all site BMD levels [23].In our study we found disease duration and age are independent risk factors for BMD and vertebral fractures In two large-scale case-control studies drawn from a population database, increase dosteo porosis incidence was found in the thyroid cancer population, but fracture incidence was not higher in the patient population compared to the control group, indicating that TSH suppression has a negative effect on bone [4,7].

Endogen hyperthyroidism was shownin large-scale cohort studies to have detrimental effects on bone remodeling, accelerate bone turnover, and reduce BMD [24].The effect of iatrogenic hyperthyroidism with TSH suppression treatment on osteoporosis is still unclear. A recent meta-analysis indicates that subclinical hyperthyroidism is associated with an elevated risk of hip and spine fractures [25].In contrast to previous findings, this study revealed that TSH levels in patients with vertebral fractures were statistically higher (p=0.0082). Additionally, the LT4 doses (p=0.079) and doses per kilogram (p=0.09) were comparable between patients with and without fractures. This result may be related to a cross-sectional study design.

The current study possesses significant strengths, having been conducted with a large cohort of DTC patients in a multicenter setting, simultaneously evaluating vertebral X-rays and BMD.

This study has several limitations. Vertebral fractures are assessed using X-ray imaging. Vertebral X-ray data were accessible for 48% of the research participants. The assessment of vertebral fractures using DXA is preferred for evaluating such fractures; however, it was unavailable in Turkey. Patients were not assessed for vitamin D levels or the use of anti-osteoporotic medications.

The study design was cross-sectional, and there were no age- and sex-matched healthy control groups but TSH mean level was calculated for each patient with current, 6 months and 1 year ago TSH levels. The exact number of vertebral fractures in subjects not receiving LT4 suppression is unknown. A retrospective study indicated a 20.8% vertebral fracture rate in osteoporotic Turkish women [26], while a cross-sectional study found a 12.1% prevalence of vertebral compression fractures in Turkish women population [27].

This study indicates that the prevalence of vertebral fractures in patients with differentiated thyroid cancer (DTC) undergoing levothyroxine suppression is higher than that reported in postmenopausal osteoporotic groups in the literature.

In conclusion findings of this study suggest that DTC patients undergoing TSH suppression have a significant risk of vertebral fractures. The incidence of vertebral fractures correlates with age, disease duration, and bone mineral density in the studied cohort. The incidence of VF was elevated in the group with severe TSH suppression.

Assessment of vertebral fractures should be integrated into the routine care of DTC survivors to ensure long-term skeletal safety.

## Data Availability

All data produced in the present study are available upon reasonable request to the authors

## Acknowledgement

We would like to thank all the participants in this study.

## Data availability

All datasets generated during and/or analyzed during the current study are not publicly available but are available from the corresponding author on reasonable request.

## Author Contribution

Dilek Gogas Yavuz led the study’s methodology and supervision and reviewed the manuscript. Ceyda Dincer Yazan carried out the data conceptualization, data analysis, and manuscript writing. Can Ilgın contributed to the data analysis. Onur Bugdaycı evaluated and reviewed the vertebral X-rays. All other authors contributed to the data conceptualization.

